# Co-occurring Pathogenic Variants in 6q27 Associated with Dementia Spectrum Disorders in a Peruvian Family

**DOI:** 10.1101/2022.11.17.22282341

**Authors:** Karla Lucia F. Alvarez, Jorge A. Aguilar-Pineda, Michelle M. Ortiz-Manrique, Marluve F. Paredes-Calderon, Bryan C. Cardenas-Quispe, Karin J. Vera-Lopez, Luis D. Goyzueta-Mamani, Miguel A. Chavez-Fumagalli, Gonzalo Davila Del-Carpio, Antero Peralta-Mestas, Patricia L. Musolino, Christian L. Lino Cardenas

**Author notes:** **Correspondence:** Christian L. Lino Cardenas.

## Abstract

Evidence suggests that there may be racial differences in risk factors associated with the development of Alzheimer’s disease and related dementias (ADRD). We used whole genome sequencing analysis and identified a novel combination of three pathogenic variants in the heterozygous state (*UNC93A*: rs7739897 and *WDR27*: rs61740334; rs3800544) in a Peruvian family with a strong clinical history of ADRD. Notably, the combination of these variants was present in two generations of affected individuals but absent in healthy members within the family. *In silico* and *in vitro* studies have provided insights into the pathogenicity of these variants. These studies predict the loss of function of the mutant UNC93A and WDR27 proteins which induced dramatic changes in the global transcriptomic signature of brain cells, including neurons, astrocytes, and especially pericytes and vascular smooth muscle cells, and thus indicating that the combination of these three variants may affect the neurovascular unit. In addition, key known molecular pathways associated with ADRD were enriched in brain cells with low levels of UNC93A and WDR27. Our findings have thus identified a genetic risk factor for familial ADRD in a Peruvian family with an Amerindian ancestral background.

## Introduction

Neurological disorders are an important cause of disability and death around the world. Interestingly, Alzheimer’s disease, dementia, Parkinson’s disease, epilepsy, schizophrenia, and autism spectrum disorder can share common anatomical alterations and cognitive defects (1). Certainly, Alzheimer’s disease is the main cause of dementia, contributes 50 to 75% of cases (2), and can be presented in two forms as defined by age: early-onset Alzheimer’s disease (EOAD), which occurs prior to 65 years of age, and late-onset Alzheimer’s disease (LOAD), which is mostly present after 65 years of age (3). Both EOAD and LOAD can have a family origin, and involve an inheritance mode of autosomal dominant transmission (4). Genetic variations in three genes, including the amyloid precursor protein (APP), presenilin 1 and 2 (PSEN1 and PSEN2), are nearly 100% penetrant, and were identified as causative of EOAD (5). On the other hand, expression of the apolipoprotein E (APOE) ε4 gene is the major risk factor for LOAD in the Caucasian population (6). There has been evidence, however, that the risk of developing Alzheimer’s disease in ε4 carriers differs among ethnic groups. For instance, ε4 carriers of African descent have a low risk of Alzheimer’s disease (7), while Amerindian genetic ancestry seemed to be protected from cognitive decline (8). Similarly, variants in the TREM2 (R47H, H157Y, and L211P) genes, which are closely associated with Alzheimer’s disease in the Caucasian population (9), were not replicated in Japanese descendants (10). These epidemiological observations indicate that genetic risk factors for neurological disorders have different effects between ethnicities.

In recent years, genome wide association studies (GWAS) have permitted the identification and characterization of multiple genetic rick loci associated with Alzheimer’s disease and related dementias (ADRD) (11). Most of these genetic loci were not associated with the APP processing, however, but rather with the immune response (TREM2, CLU, CR1, CD33, EPHA1, MS4A4A/MS4A6E), endosomal trafficking (PICALM, BIN1, CD2AP) or lipid metabolism (ABCA7) (12). An overlap between Alzheimer’s disease pathogenic variants and other neurogenerative or neuropsychiatric disorders has also been reported, indicating a shared genetic and molecular origin. For example, an Alzheimer’s disease variant in the TREM2 gene (rs75932628) was also correlated with amyotrophic lateral sclerosis (13), while a variant in the MARK2 gene (rs10792421) was associated with Alzheimer’s disease and bipolar disorder (14).

Identifying individuals at high risk of ADRD remains a global health need, and a major challenge for minority populations. Here, we performed a whole genome sequencing (WGS) analysis for a Peruvian family with a strong clinical history of ADRD, including Alzheimer’s disease, schizophrenia, and cognitive deficit. We also explored the effect of these variants on the neurovascular unit of the brain through *in silico* and *in vitro* studies.

## Materials and methods

### Patient sample collection

A family (n=14) originally from Peru, with five members diagnosed with neurological and neuropsychiatry disorders, was enrolled in this study. Non-familial AD patients (n=8) and healthy individuals (n=50) were recruited for the variant validation study. The selection criteria for the healthy individuals was as follows: age >60 years, without signs of dementia, and no familial history of AD. Probable AD was diagnosed according to the guidelines of the National Institute of Neurological and Communicative Disorders and the Stroke and Alzheimer Disease and Related Disorders Association (15). Whenever possible, the cognitive status of each family member was diagnosed based on a neuropsychological test (MoCa blind test and the clock drawing test). The cutoff for a normal MoCA score was 18, and for a normal clock drawing test was 6.

### Genetic Analysis

Genomic DNA was extracted from saliva samples using the prepIT.L2P reagent (Genotek, Cat. No PT-L2P-5) according to the manufacturer’s instructions. The qualifying genomic DNA samples were randomly fragmented using Covaris Technology, obtaining a fragment of 350bp. The DNA nanoballs (DNBs) were produced using rolling circle amplification (RCA), and the qualified DNBs were loaded into the patterned nanoarrays. The WGS was conducted on the BGISEQ-500 platform (BGI Genomics, Shenzhen, China). Raw sequencing reads were aligned to the human reference genome (GRCh38/HG38) with the Burrows-Wheeler Aligner (BWA) software, and variant calling was performed with the Genome Analysis Toolkit (GATK v3.5) according to best practice. On average, 88.10% mapped successfully and 93.23% mapped uniquely. The duplicate reads were removed from total mapped reads, resulting in a duplicate rate of 2.48%, and a 30.72-fold mean sequencing depth on the whole genome, excluding gap regions. On average per sequencing individual, 99.34% of the whole genome, excluding gap regions, were covered by at least 1X coverage, 98.78% had at least 4X coverage and 97.42% had at least 10X coverage.

### Sanger sequencing

Based on the results of the WGS, the variants that were present in affected members of the family but absent in healthy individuals were validated using Sanger sequencing. All PCR products were sequenced using an ABI 3130 Genetic Analyzer. Sequence analysis was performed with the Chromas program in the DNASTAR analysis package. The PCR and sequencing primers are shown in Table 1.

**Table 1.**
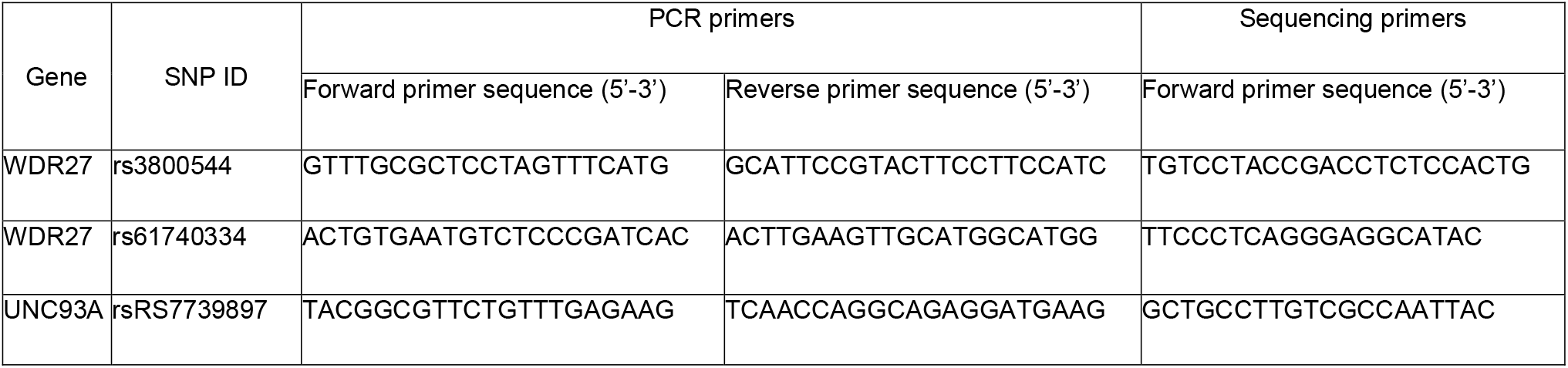
PCR and sequencing primers

### Cell lines

Primary human vascular smooth muscle cells (VSMCs) from carotid of healthy donors were purchased from Cell Applications Inc. (Cat. No 3514k-05a, neural crest origin). Human brain vascular pericytes were purchased from ScienCell (Cat. No 1200). Human neurons (SH-SY5Y) were purchased from ATCC (Cat. No CRL-2266). Human astrocytes were purchased from Cell Applications Inc. (Cat. No 882A05f).

### RNA sequencing and qPCR

Total RNA was extracted using a miRNeasy kit (Qiagen, Cat. No 217084) following the manufacturer’s protocol instructions. The BGISEQ platform was used for RNA-seq, generating some 4.28G Gb bases per sample, on average. The average mapping ratio with the reference genome was 97.01%, and the average mapping ratio with gene was 74.05%; 17029 genes were identified. We used HISAT to align the clean reads to the reference genome, and Bowtie2 to align the clean reads to the reference genes. 100 ng of total RNA was used for qPCR, as the starting template for cDNA synthesis. The cDNA was prepared by reverse transcription (RT), and gene expression was analyzed by quantitative PCR (qPCR) on a SYBR green system (Applied Biosystems). Expression results were analyzed using the DDCT method, and GAPDH (encoding glyceraldehyde-3-phosphate dehydrogenase) was used as a house-keeping gene. Fold changes were calculated as the average relative to the control carotid as the baseline.

### Computational Details

#### System building, structural refinements and molecular dynamic simulations (MDS)

The Q86WB7-1 (UNC93A, 457 aa), and A2RRH5-4 (WDR27, 827 aa) sequences (16,17) were used to build the 3D wild-type protein structures using the I-TASSER server (18). The mutant variants (UNC93A V409I, and WDR27 R467H - T542S), were built based on these 3D models by performing site direct mutagenesis using UCSF Chimera software (19). In order to avoid the residue overlapping in all protein systems, a structural refinement was carried out using ModRefiner server (20). Classical MD simulations were performed using the GROMACS 2020.4 package with the OPLS-AA force field parameters (21,22). All protein systems were built in a triclinic simulation box with periodic boundary conditions (PBC) in all directions (x, y, and z). They were then solvated using the TIP4P water model (23), and Cl^-^ or Na^+^ ions were used to neutralize the total charge in the simulation box. The mimicking of physiological conditions was performed by ionic strengthening, with the addition of 150 mM NaCl. The distance of the protein surfaces to the edge of the periodic box was set at 1.5 nm, and 1 fs step was applied to calculate the motion equations using the Leap-Frog integrator (24). The temperature for proteins and water-ions in all simulations was set at 309.65 K using the modified Berendsen thermostat (V-rescale algorithm), with a coupling constant of 0.1 ps (25). The pressure was maintained at 1 bar using the Parrinello-Rahman barostat with compressibility of 4.5×10^−5^ bar^-1^ and coupling constant of 2.0 ps (26). The particle mesh Ewald method was applied to long-range electrostatic interactions with a cutoff equal to 1.1 nm for non-bonded interactions, with a tolerance of 1×10^5^ for contribution in the real space of the 3D structures. The Verlet neighbor searching cutoff scheme was applied with a neighbor-list update frequency of 10 steps (20 fs). Bonds involving hydrogen atoms were constrained using the linear constraint solver (LINCS) algorithm (27). Energy was minimized in all simulations with the steepest descent algorithm for a maximum of 100,000 steps. We performed two steps for the equilibration process; one step of dynamics (1 ns) in the NVT (isothermal-isochoric) ensemble, followed by 2 ns of dynamics in the NPT (isothermal-isobaric) ensemble. The final simulation was then performed in the NPT ensemble for 500 ns followed by the analysis of the structures and their energy properties.

#### Structural and energetic analysis of 3D protein structures

All MD trajectories were corrected, and the 3D structures were recentered in the simulation boxes. RMSD, RMSF, radius of gyrations, H-bonds, residue distances, and solvent accessible surface area analyses were performed using the Gromacs tools, and the results were plotted using XMGrace software. We used the UCSF Chimera, VMD software packages to visualize the structures. Atomic interactions and 2D plots were analyzed using the LigPlot software packages (28). Electrostatic potential (ESP) surfaces were calculated using the APBS (Adaptive Poisson-Boltzmann Solver) software, and the PDB2PQR software was used to assign the charges and radii to protein atoms (29).

#### Calculation of binding free energy

The Molecular Mechanics Poisson-Boltzmann Surface Area (MM/PBSA) of free energies and energy contribution by individual residues was calculated to analyze the effect of amino-acid substitutions on the different structures using the last 100ns of MD trajectories and the g_mmpbsa package (30). The interacting energy was calculated using the following equation:

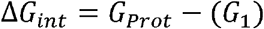

where the term G1 is the free energy of the different sites of the protein, and GProt is the free energy of entire 3D structure. In this context, the free energy of each term was calculated as follows:

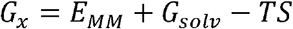

where EMM is the standard mechanical energy (MM) produced from bonded interactions, electrostatic interactions, and van der Waals interactions. Gsolv, is the solvation energy that includes the free energy contributions of the polar and nonpolar terms. The TS term refers to the entropic contribution, and was not included in this calculation due to the computational costs (30,31). Finally, 309 Kelvin (K) of temperature was used as the default parameter in all our calculations.

## Results

### Genetic analyses

Genealogic investigations allowed us to identify five members of a Peruvian family with a strong clinical history of ADRD, including dementia, Alzheimer’s disease and schizophrenia across two generations. In order to detect genetic risk factors associated with the development of the neurologic disorders observed in this family, we performed a WGS analysis on affected and healthy members (n=14) of the family. By using the BGISEQ-500 platform, we obtained an average of 113,895.38 Mb of raw bases. After removing low-quality reads, we obtained an average of 106,676.25 Mb clean reads, identifying a total of 3,933,470 SNPs. We then selected coding variants that met the following two criteria: first, candidate variants that harbored at least one “disruptive” or missense variant, and second, variants that were present in affected probands, but not in unaffected members of the family. We identified three coding variants that segregated across two generations of affected individuals. These variants were found to be located at chr6:167728791 (*UNC93A*; rs7739897), at chr6:170047902 (*WDR27*; rs61740334), and at chr6:170058374 (*WDR27*; rs3800544) (Fig. 1A-B). Two different Sanger PCR sequencing platforms were used to validate the presence of these SNPs (Fig. 1C). Several studies have also found SNPs in genes located on chromosome 6q with a significant connection to neurological diseases (32,33), however, the *UNC93A* and *WDR27* variants have not yet been associated with ADRD. It is worthy of note that none of the currently known Alzheimer’s disease-associated variants were present in this Peruvian family.

**Figure 1.**
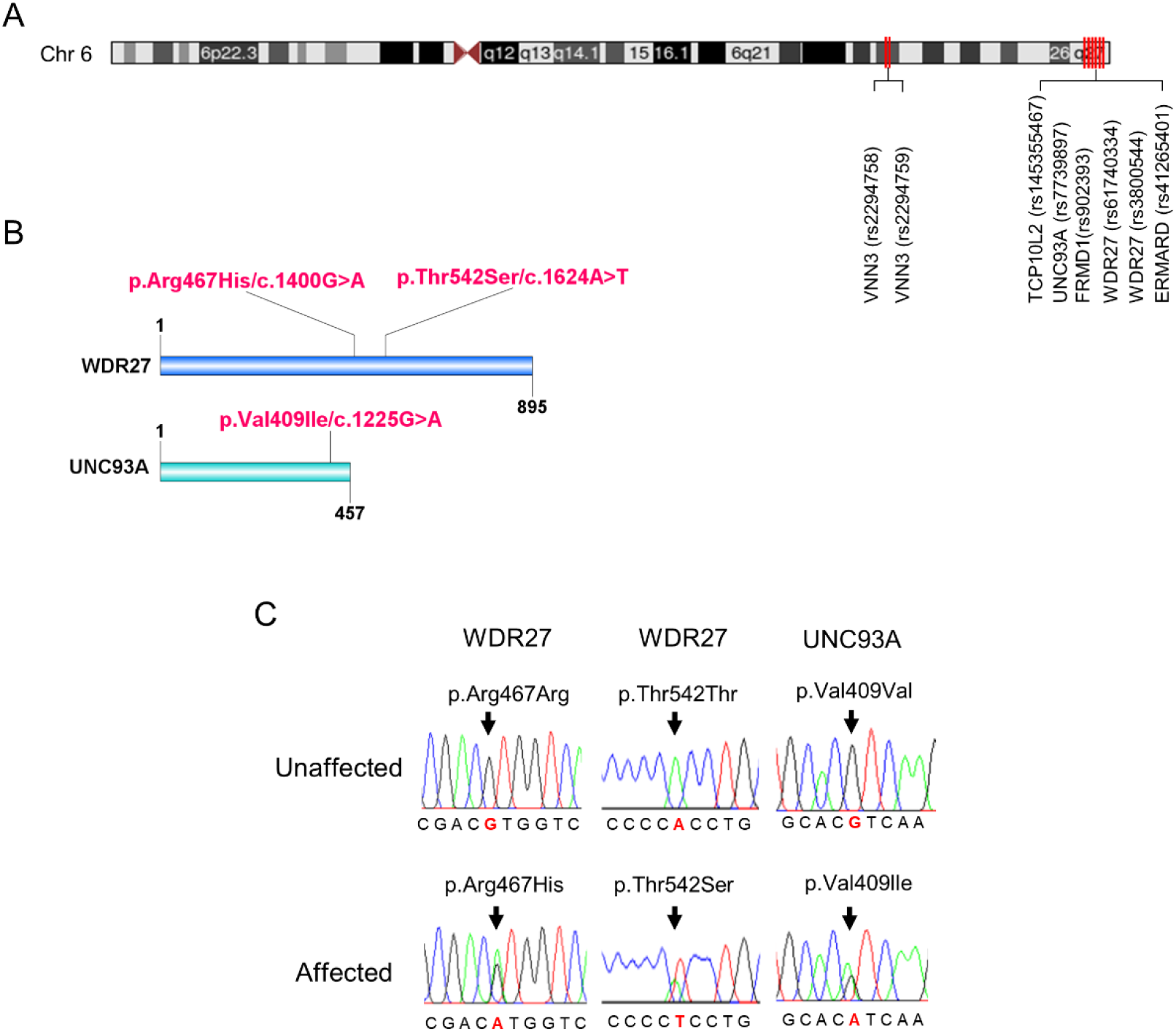
Variants identified in family affected members. (A) Chromosomal position of UNC93A and WDR27 genes. (B) Location of a mutation in the protein structure. (C) An electropherogram from affected probands showing a base pair change in the UNC93A gene (Val409Ile/c.1225G>A) and in the WDR27 gene (Thr542Ser/c.1624A>T; Arg467His/c.1400G>A), compared to healthy controls.

In order to further confirm an association between the *UNC93A* and *WDR27* variants and familial genetics risk for neurologic disorders, we analyzed their presence in unrelated healthy Peruvians (n=50) and unrelated individuals with neurological disorders (probable Alzheimer’s disease, n=8). As shown in Table 2, the UNC93A variant (V409I) was present in 1/50 of the healthy group, and the WDR27 variants (Thr542S and Arg467His) were present in 2/50 of the healthy group. Interestingly, the three variants did not co-exist in any healthy individuals and were absent in volunteers diagnosed with probable Alzheimer’s disease with no familial history of ADRD. These findings suggest the co-occurrence of these three variants may be related to neurological disorders in a Peruvian family.

**Table 2.** Variants in unrelated volunteers.

| Gene | Variant | Reference SNP number | Unrelated Controls (n=50) | Unrelated volunteers with ADRD (n=8) |
| --- | --- | --- | --- | --- |
| UNC93A | V409I | rs7739897 | 1 | 0 |
| WDR27 | Thr542S | rs61740334 | 2 | 0 |
| WDR27 | Arg467His | rs3800544 | 2 | 0 |

SNP: single nucleotide polymorphism; rs: reference sequence

### Structural analysis of the WDR27 and UNC93A variants

UNC93A genes encode a transmembrane protein (457 amino acids) that has 11 alpha-helices and is mainly expressed in the brain, kidney, and liver (34). The WDR27 gene encodes a scaffold protein with multiple WD repeat domains and is ubiquitously expressed in the human body. We used *in silico* approaches to provide insights into the molecular and structural effect of the UNC93A (V409I) and WDR27 (Arg467His and T542S) variants associated with familial ADRD. We therefore built the human structure of the UNC93A and WDR27 protein by homology modelling and performed a site direct mutagenesis to generate the mutated proteins using the UCSF Chimera software (Fig. 2A-B). Molecular dynamics simulations (MDS) for 500 ns were performed to stabilize the physical motions of atoms in both proteins, to mimic physiological conditions. Importantly, we observed that at the beginning of the MDS (0-50 ns) the residue V409 of the wild-type UNC93 protein interacts with the lipid bilayer of the cell membrane. After 500ns of MDS the V409 is internalized toward to the protein active transmembrane conduct region in which the subsequent interactions with ligands or ionic exchanges occur. These movements were characterized by high structural vibration and epitope exposure of the UNC93A’s ectodomain (aa 200-300) of the protein towards the surface of cell membrane (Fig. 2C). In contrast, the mutant I409 blocks the internalization of this residue and loses its capacity to move towards to the protein active transmembrane conduct region due to the loss of about 50% of the global residual vibration (Fig. 2D). As a result of this change in the amino acid, the UNC93A loses its capacity for ion exchange and interaction with potential ligands or partners. Regarding the WDR27 protein, both His467 and S542 variants induce the internalization of these residues in the part of the hinge domain of the protein predicting the loss of function and capacity to interact with other partners (Fig. 2E). We also observed that the wild-type and mutant proteins are very stable due their closely residual vibration and epitope exposure patterns (Fig. 2F). The solvent accessible surface area (SASA) analysis demonstrated that the UNC93A (I409) protein increased the surface area of the mutated amino acid and its environment (Fig. 2G), while the WDR27 variants reduced its SASA (Fig. 2H). The effect on the amino acid substitutions for both proteins was determined using the Molecular Mechanics Poisson-Boltzmann Surface Area (MM/PBSA) calculation of free energies and energy contribution using the last 150ns of MD trajectories. Remarkably the

**Figure 2.**
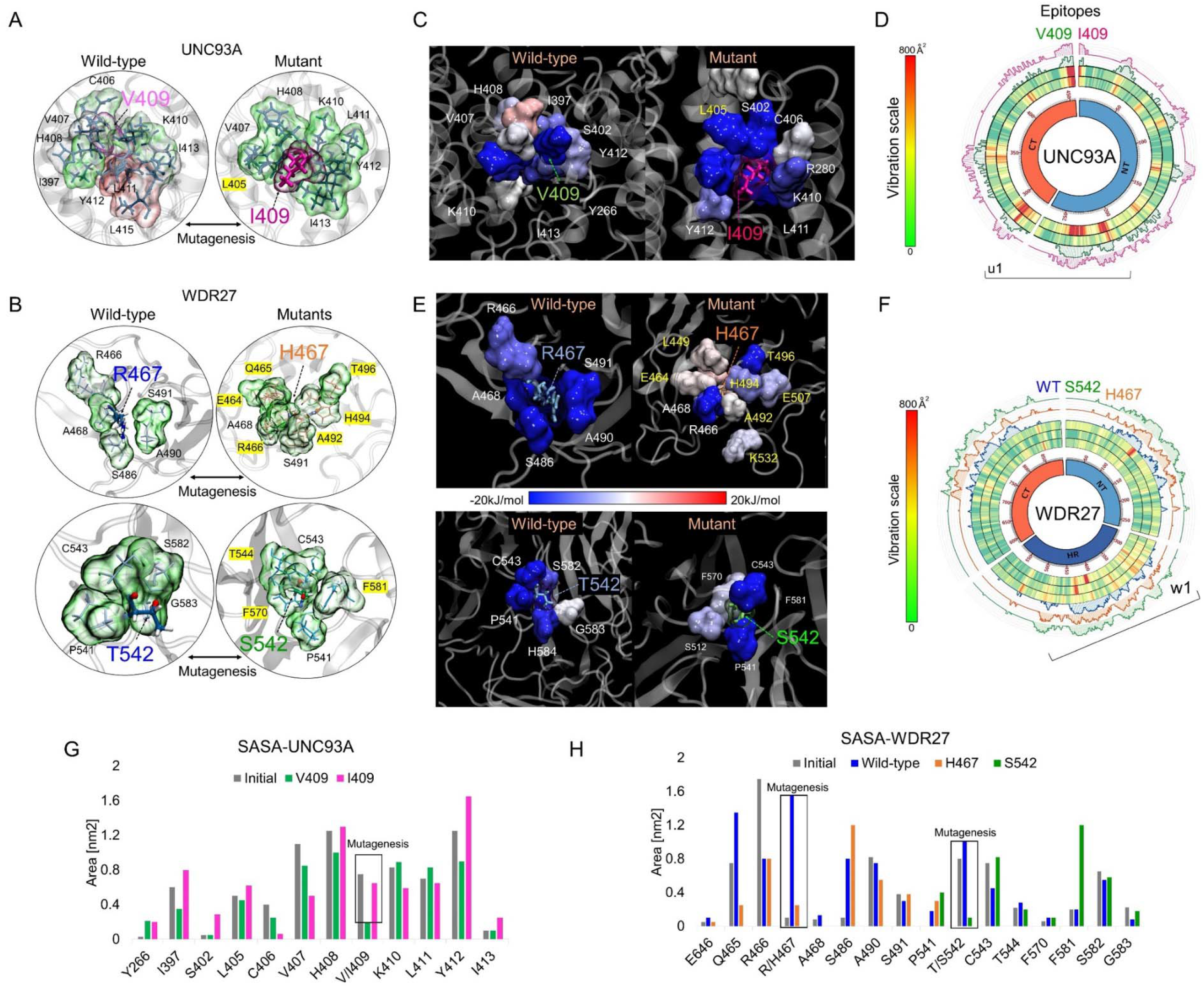
*In silico* analysis of UNC93A and WDR27 amino acid substitutions predicts loss of protein function. (A and B) show the UNC93A and WDR27 wild-type and mutant proteins. (C and E) show the MM-PBSA calculation of main energetic interactions of residues at mutation sites. Blue indicates favorable energies and red unfavorable energies. (D and F) show a Circos plot of the wild-type and mutant full-length protein. The heat map represents the vibrational movement of each residue throughout MD simulations at the scale bar values. The outer histograms show the regions most likely (>50%) to be epitopes. (G and H) show the solvent-accessible surface area (SASA) average values of the wild-type and mutation residues and their neighbors.

I409 increased the affinity of the protein to the membrane, indicating a reduced capacity for internalization, while the WDR27 variants reduced the protein stabilization of both His467 and S542 mutated amino acids. Together these findings indicate that the UNC93A and WDR27 variants have a strong effect on the functionality and ability to interact with their environment, and thus potentially affect brain homeostasis.

### Functional analysis of loss of WDR27 and UNC93A gene expression *in vitro*

We next investigated how the co-occurring inhibition of *UNC93A* and *WDR27* gene expression affects the cellular homeostasis of brain cell lines, including neurons, pericytes, astrocytes and VSMCs. In order to achieve this goal, we simultaneously silenced both *UNC93A* and *WDR27* genes using a siRNA approach and performed a high throughput RNA sequencing. Differential expression analysis demonstrates that VSMCs had the highest numbers of differentially expressed genes (DEG=2231), followed by pericytes (DEG=2091), while astrocytes (DEG=226) and neurons (DEG=191) showed a modest change in gene expression compared with control groups (Fig. 3A), suggesting that mutations promoting a loss of function of the *UNC93A* and *WDR27* genes affect the neurovascular unit of the brain. Pathway analysis of VSMC and pericyte gene signatures identified enrichment for multiple known molecular pathways associated with neurological disorders, such as impaired autophagy pathways signaling (35), ubiquitin-mediated proteolysis (36), unproductive metabolism signaling (37) inflammation and necroptosis (38) (Fig. 3B), while neurons showed a gene signature enriched for cellular senescence, cytokine-cytokine interactions and apoptosis. Astrocytes showed only modest enrichment for necroptosis and the proteasomal degradation pathway (Fig. 3B). Similarly to our results, previous *in vivo* studies have reported a direct connection between autophagy activation and UNC93A levels in the healthy brains of mice under starvation, indicating the potential role of UNC93A in metabolic stability, energy uptake and nutrient transport in the brain (34). Interestingly several neurological diseases have been associated with defects of autophagy and metabolism homeostasis, including Alzheimer’s, Parkinson’s, and Huntington’s diseases (39–41). We also observed that multiple genes previously related to Alzheimer’s disease, such as CLU (42) SQSTM1 (43) GPC6 (44) and ABCA7 (45), were dysregulated in brain cells deficient in UNC93A and WDR27 (Fig. 3C). Interestingly, the strongest genetic risk factor for Alzheimer’s disease, APOE, was also modulated in pericytes and neurons (Fig. 3C).

**Figure 3.**
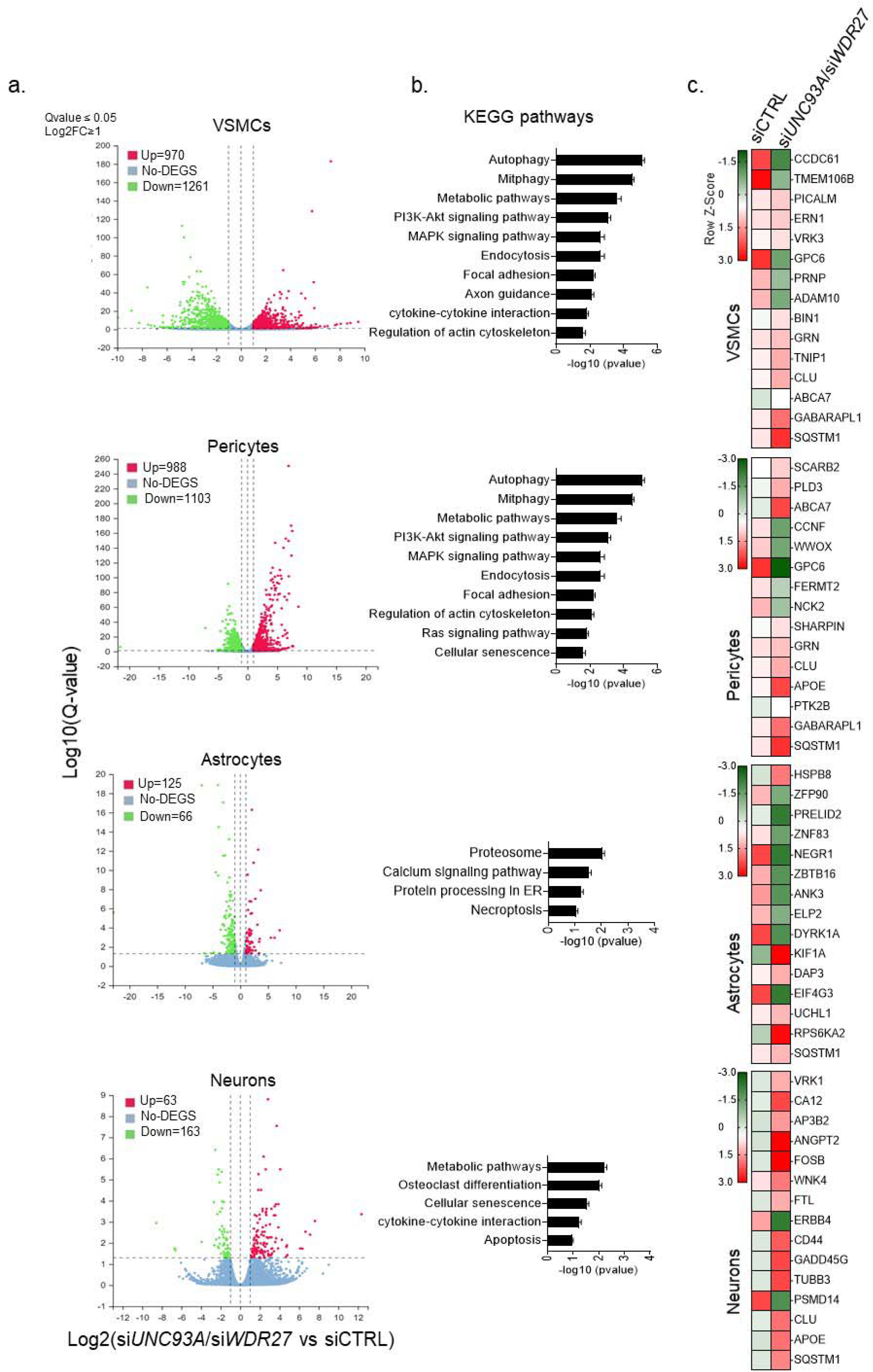
Loss of UNC93A and WDR27 affect the global transcriptomic signature of brain cell lines *in vitro*. (A) Volcano plot of dysregulated genes in four different brain cell types. (B) Shows the KEGG pathways analysis. (C) Heatmap of dysregulated genes previously associated with neurodegenerative disease.

## Discussion

Recent studies have shown racial disparities in ADRD diagnosis between white and minority groups (46,47). Diverse evidence suggests that there may be racial differences in risk factors associated with the development of ADRD (48–50). Risk factors such as genetics, age, lifestyle, and co-morbid cardiovascular disease can be useful to understand the incidence, prevalence, and predisposition of an individual to ADRD. Despite the research progress on racial differences in ADRD in developed countries, the diagnosis of ADRD in developing countries (e.g., Asian, African, and South American countries) deserves more recognition for its contribution to the global burden of Alzheimer’s disease (51). The limited resources to address mental health issues, the lack of adequate technology to diagnose ADRD, and few funding agencies that support research studies are also major challenges faced by public health systems in developing countries. According to the latest WHO report in 2018, ADRD deaths in Peru reached an astonishing 5.20% of total deaths (52). Notably the Peruvian population has a strong Amerindian ancestral background (approximately 80%), compared to other Latin American populations (53,54), indicating an opportunity to identify ancestry-specific genetic modifiers associated with the development of ADRD.

Here we report an inheritance risk factor for ADRD in a Peruvian family with Amerindian ancestral background. We identified a novel combination of three pathogenic variants in the heterozygous state (*UNC93A*: rs7739897 and *WDR27*: rs61740334; rs3800544) which segregated across two generations in a family with a strong clinical history of ADRD. Notably, the combination of these variants was present in members with neurological disorders but absent in healthy individuals. Importantly, although these three SNPs are fairly common in European American and African American ancestry populations (MAF = 1.78 - 17.09) (https://evs.gs.washington.edu/EVS/) the combined effect of these variants was not previously studied. Our findings thus suggest that the combination of these variants is necessary to manifest the disease. Supporting our hypothesis, it has been reported variants that have no impact on health when found individually but cause severe disease when in combination with other genetic variants (55).

Our *in silico* analysis of the 3D structure of the mutant UNC93A (V409I) and WDR27 (Arg467His and T542S) proteins demonstrates that changes in the amino acid sequences have a dramatic effect on the conformational structure, predicting the loss of function of both proteins. The exact biological role of the UNC93A protein remains unknown, however. For instance, some studies have identified the potential role of UNC93A as solute carrier, and in ion homeostasis (56). Its expression seemed to be associated with increased metabolic activity in organs such as the brain and kidneys (34). In this context, we observed that amino acid substitution (I409) in the UNC93A protein reduced its capacity for ion exchange and interaction with potential ligands or partners, indicating a negative effect on UNC93A bioactivity in the brain. Our *in silico* analysis of the mutant WDR27 proteins demonstrated that both amino acid substitutions induced the internalization of the hinge domain of the protein, affecting its segmental flexibility and ability to clamp down on its substrates or ligands. Similarly, little is known about the WDR27 biological functions in the brain, however, an SNP in the intergenic region adjoining WDR27 (rs924043) was associated with type 1 diabetes (57), and its duplication has been seen in obese patients (58). These results suggest the involvement of UNC93A and WRD27 in metabolic syndrome and related diseases.

Importantly, the brain is the most complex and metabolically active organ, being equipped with a sophisticated network of specialized cell types such as neurons, microglia, astrocytes, pericytes, and VSMCs. In recent years, diverse studies have demonstrated the contribution of these cells to Alzheimer’s disease pathogenesis (59,60). In order to investigate the potential effects of a loss of function of UNC93A and WDR27, we used gene silencing technologies to simultaneously reduce the expression of both proteins in four brain cell types to mimic the clinical phenotype of members of the family with ADRD. Our KEGG pathway enrichment analysis showed that autophagy, mitophagy and metabolic pathways are the most affected in both UNC93A and WDR27 inhibitory conditions. Interestingly, these pathways play an important role in Aβ clearance, and thus its dysfunction may lead to the development Alzheimer’s disease (61). As reduced autophagy activity was related to increasing cell death in response to intracellular stress (62), these variants could have a negative effect on the BBB integrity.

Our study has several limitations. First, a lack of access to imagological studies meant that we were not able to correlate the variants with damaged areas of the brain, however, the MoCa test corroborated that brain areas associated with cognitive domains, predominantly temporal and frontal lobe areas, are damaged. Second, we could not find a validation family for the combination of these variants, However, it is possible that these combinatory variants are only present in the reported family. This could be a similar case to that reported for the PSEN1 (E280A) mutation, which only affects the Colombian family descendant of a Spanish conquistador (63), or the mutation in the PSEN2 gene (N141I) that is only present in families with German descendants who emigrated to a southern Volga region in Russia in the 1760s (64). Despite these limitations, this study reports for the first time a new genetic rick loci associated with ADRD, and the great importance of the UNC93A and WDR27 genes in brain biology.

## Data Availability

All data produced in the present study are available upon reasonable request to the authors

## Availability of data and materials

All raw data is stored in cloud storage and will be made available upon request.

## Ethics statement

This study was approved by the Ethics Committee for Research of the EsSalud Hospital (CIEI) of Arequipa-Peru. Saliva samples were obtained from participants who had previously given informed consent.

## Author contributions

CLLC conceived the work with KLFA and GDDC. CLLC and KLFA designed the work. CLLC and KLFA performed the experiments. KLFA, MMOM, LDGM, MACF, and KJVL collected the samples. JAAP performed the *in silico* analysis. MFPC performed and analyzed the neurological tests. PLM analyzed the neurological tests. CLLC and KLFA analyzed and interpreted the data of the study. APM provided the clinical samples, and made a substantial contribution to the design of the study. GDDC and CLLC supervised the study. CLLC and KLFA wrote the paper. All authors read and approved the final manuscript.

## Acknowledgments

We are very grateful to all the volunteers who participated in this study. In addition, we thank Drs. Claudia Caracela-Zeballos, Jessica L. Lewis-Paredes, Juan M Pacheco-Salazar, Froilan Huaraya, Rita Montesinos-Nieto, and Badhin Gomez for their contribution to this study.

## References

1. Kochunov P, Zavaliangos-Petropulu A, Jahanshad N, Thompson PM, Ryan MC, Chiappelli J, et al. A white matter connection of schizophrenia and Alzheimer’s Disease. Schizophr Bull. 2021;47(1):197–206.

2. Report AA. 2021 Alzheimer’s disease facts and figures. Alzheimer’s Dement. 2021;17(3):327–406.

3. Mendez MF. Early-onset Alzheimer Disease. Neurol Clin. 2017;35(2):263–81.

4. Gatz M, Reynolds CA, Fratiglioni L, Johansson B, Mortimer JA, Berg S, et al. Role of genes and environments for explaining Alzheimer disease. Arch Gen Psychiatry. 2006;63:168–74.

5. Van Cauwenberghe C, Van Broeckhoven C, Sleegers K. The genetic landscape of Alzheimer disease: Clinical implications and perspectives. Genet Med. 2016;18(5):421–30.

6. Guerreiro RJ, Gustafson DR, Hardy J. The genetic architecture of Alzheimer’s disease: Beyond APP, PSENS and APOE. Neurobiol Aging. 2012;33(3):437–56.

7. Farrer LA, Cupples LA, Haines JL, Hyman B, Kukull WA, Mayeux R, et al. Effects of age, sex, and ethnicity on the association between apolipoprotein E genotype and Alzheimer disease: A meta-analysis. J Am Med Assoc. 1997;278(16):1349–56.

8. Granot-Hershkovitz E, Tarraf W, Kurniansyah N, Daviglus M, Isasi CR, Kaplan R, et al. APOE alleles’ association with cognitive function differs across Hispanic/Latino groups and genetic ancestry in the study of Latinos-investigation of neurocognitive aging (HCHS/SOL). Alzheimer’s Dement. 2021;17:466–74.

9. Guerreiro R, Wojtas A, Bras J, Carrasquillo M, Rogaeva E, Majounie E, et al. TREM2 variants in Alzheimer’s Disease. N Engl J Med. 2013;368(2):117–27.

10. Miyashita A, Wen Y, Kitamura N, Matsubara E, Kawarabayashi T, Shoji M, et al. Lack of genetic association between TREM2 and late-onset Alzheimer’s disease in a Japanese population. J Alzheimer’s Dis. 2014;41(4):1031–8.

11. Kunkle BW, Grenier-Boley B, Sims R, Bis JC, Damotte V, Naj AC, et al. Genetic meta-analysis of diagnosed Alzheimer’s disease identifies new risk loci and implicates Aβ, tau, immunity and lipid processing. Nat Genet. 2019;51(3):414–30.

12. Bellenguez C, Grenier-Boley B, Lambert JC. Genetics of Alzheimer’s disease: where we are, and where we are going. Curr Opin Neurobiol. 2020;61:40–8.

13. Cady J, Koval ED, Benitez BA, Zaidman C, Jockel-balsarotti J, Allred P, et al. The TREM2 variant p.R47H is a risk factor for sporadic amyotrophic lateral sclerosis. 2014;71(4):449–53.

14. Drange OK, Bjerkehagen Smeland O, Shadrin AA, Finseth PI, Witoelar A, Frei O, et al. Genetic overlap between alzheimer’s disease and bipolar disorder implicates the MARK2 and VAC14 genes. Front Neurosci. 2019;13(220):1–11.

15. McKhann G, Knopman DS, Chertkow H, Hyman BT, Jr CRJ, Kawas CH, et al. The diagnosis of dementia due to Alzheimer’s disease: Recommendations from the National Institute on Aging-Alzheimer’s Association workgroups on diagnostic guidelines for Alzheimer’s disease. Alzheimers Dement. 2011;7(3):263–9.

16. WDR27 - WD repeat-containing protein 27 - Homo sapiens (Human) [Internet]. [cited 2020 Jun 10]. Available from: https://www.uniprot.org/uniprot/A2RRH5#A2RRH5-4

17. UNC119 - Protein unc-119 homolog A - Homo sapiens (Human) [Internet]. [cited 2020 Jun 10]. Available from: https://www.uniprot.org/uniprot/Q86WB7#Q86WB7-1

18. Zheng W, Zhang C, Li Y, Pearce R, Bell EW, Zhang Y. Folding non-homologous proteins by coupling deep-learning contact maps with I-TASSER assembly simulations. Cell Reports Methods [Internet]. 2021;1:100014.

19. Pettersen EF, Goddard TD, Huang CC, Couch GS, Greenblatt DM, Meng EC, et al. UCSF Chimera - A visualization system for exploratory research and analysis. J Comput Chem. 2004;25(13):1605–12.

20. Xu D, Zhang Y. Improving the physical realism and structural accuracy of protein models by a two-step atomic-level energy minimization. Biophys J [Internet]. 2011;101(10):2525–34.

21. Kutzner C, Páll S, Fechner M, Esztermann A, de Groot BL, Grubmüller H. More bang for your buck: Improved use of GPU nodes for GROMACS 2018. J Comput Chem. 2019;40(27):2418–31.

22. Jorgensen WL, Maxwell DS, Tirado-Rives J. Development and testing of the OPLS all-atom force field on conformational energetics and properties of organic liquids. J Am Chem Soc. 1996;118(45):11225–36.

23. Jorgensen WL, Chandrasekhar J, Madura JD, Impey RW, Klein ML. Comparison of simple potential functions for simulating liquid water. J Chem Phys. 1983;79(2):926– 35.

24. Hockney RW, Goel SP, Eastwood JW. Quiet high-resolution computer models of a plasma. J Comput Phys. 1974;14(2):148–58.

25. Berendsen HJC, Postma JPM, Van Gunsteren WF, Dinola A, Haak JR. Molecular dynamics with coupling to an external bath. J Chem Phys. 1984;81(8):3684–90.

26. Bussi G, Donadio D, Parrinello M. Canonical sampling through velocity rescaling. J Chem Phys. 2007;126(1).

27. Hess B. P-LINCS: A parallel linear constraint solver for molecular simulation. J Chem Theory Comput. 2008;4(1):116–22.

28. Wallace AC, Laskowski RA, Thornton JM. Ligplot: A program to generate schematic diagrams of protein-ligand interactions. Protein Eng Des Sel. 1995;8(2):127–34.

29. Baker NA, Sept D, Joseph S, Holst MJ, McCammon JA. Electrostatics of nanosystems: Application to microtubules and the ribosome. Proc Natl Acad Sci U S A. 2001;98(18):10037–41.

30. Kumari R, Kumar R, Lynn A. G-mmpbsa -A GROMACS tool for high-throughput MM-PBSA calculations. J Chem Inf Model. 2014;54(7):1951–62.

31. Rastelli G, Del Rio A, Gianluca D, Miriam S. Fast and accurate predictions of binding free energies using MM-PBSA and MM-GBSA. Wiley Intersci. 2010;31:797–810.

32. Kohn Y, Lerer B. Excitement and confusion on chromosome 6q: The challenges of neuropsychiatric genetics in microcosm. Mol Psychiatry. 2005;10(12):1062–73.

33. Naj AC, Beecham GW, Martin ER, Gallins PJ, Powell EH, Konidari I, et al. Dementia revealed: Novel chromosome 6 locus for Late-onset alzheimer disease provides genetic evidence for Folate-pathway abnormalities. PLoS Genet. 2010;6(9).

34. Ceder MM, Lekholm E, Hellsten S V., Perland E, Fredriksson R. The Neuronal and Peripheral Expressed Membrane-Bound UNC93A Respond to Nutrient Availability in Mice. Front Mol Neurosci. 2017;10(351).

35. Li Q, Liu Y, Sun M. Autophagy and Alzheimer’s Disease. Cell Mol Neurobiol. 2016;37(3):377–88.

36. Upadhya SC, Hegde AN. Role of the ubiquitin proteasome system in Alzheimer’s disease. BMC Biochem. 2007;8(S12).

37. Van Der Velpen V, Teav T, Gallart-Ayala H, Mehl F, Konz I, Clark C, et al. Systemic and central nervous system metabolic alterations in Alzheimer’s disease. Alzheimer’s Res Ther. 2019;11(93).

38. Zhang G, Zhang Y, Shen Y, Wang Y, Zhao M, Sun L. The potential role of ferroptosis in Alzheimer’s Disease. J Alzheimer’s Dis. 2021;907–25.

39. Xu Y, Propson NE, D. S, Xiong W, Zheng H. Autophagy deficiency modulates microglial lipid homeostasis and aggravates tau pathology and spreading. Proc Natl Acad Sci U S A. 2021;118(27):1–10.

40. Wang B, Abraham N, Gao G, Yang Q. Dysregulation of autophagy and mitochondrial function in Parkinson’s disease. Transl Neurodegener [Internet]. 2016;5(1):1–9.

41. Croce KR, Yamamoto A. A role for autophagy in Huntington’s disease Katherine. Neurobiol Dis. 2019;122:16–22.

42. Jun G, Naj AC, Beecham GW, Wang LS, Buros J, Gallins PJ, et al. Meta-analysis confirms CR1, CLU, and PICALM as Alzheimer disease risk loci and reveals interactions with APOE genotypes. Arch Neurol. 2010;67(12):1473–84.

43. Cuyvers E, van der Zee J, Bettens K, Engelborghs S, Vandenbulcke M, Robberecht C, et al. Genetic variability in SQSTM1 and risk of early-onset Alzheimer dementia: A European early-onset dementia consortium study. Neurobiol Aging. 2015;36(5):2005.e15-2005.e22.

44. Kunkle BW, Schmidt M, Klein HU, Naj AC, Hamilton-Nelson KL, Larson EB, et al. Novel Alzheimer Disease risk loci and pathways in african american individuals using the African genome resources panel: A meta-analysis. JAMA Neurol. 2021;78(1):102– 13.

45. De Roeck A, Van Broeckhoven C, Sleegers K. The role of ABCA7 in Alzheimer’s disease: evidence from genomics, transcriptomics and methylomics. Acta Neuropathol [Internet]. 2019;138(2):201–20.

46. Lennon JC, Aita SL, Bene VAD, Rhoads T, Resch ZJ, Eloi JM, et al. Black and White individuals differ in dementia prevalence, risk factors, and symptomatic presentation. Alzheimer’s Dement. 2022;18(8):1461–71.

47. Suran M. Racial Disparities in Dementia Diagnoses. JAMA - J Am Med Assoc. 2022;327(8):709.

48. Barnes LL. Alzheimer disease in African American individuals: increased incidence or not enough data? Nat Rev Neurol. 2022;18(1):56–62.

49. Mayeda Elizabeth Rose, M Maria Glymor CPQ, Whitmer RAW. Inequalities in dementia incidence between six racial and ethnic groups over 14 years. Physiol Behav. 2017;176(3):139–48.

50. Brewster P, Barnes L, Haan M, Johnson JK, Manly JJ, Nápoles AM, et al. Progress and future challenges in aging and diversity research in the United States. Alzheimers Dement. 2019;15(7):995–1003.

51. Chávez-Fumagalli MA, Shrivastava P, Aguilar-Pineda JA, Nieto-Montesinos R, Del-Carpio GD, Peralta-Mestas A, et al. Diagnosis of Alzheimer’s disease in developed and developing countries: systematic review and meta-analysis of diagnostic test accuracy. J Alzheimer’s Dis Reports. 2021;5(1):15–30.

52. World heatlh rankings [Internet]. [cited 2021 Dec 15]. Available from: https://www.worldlifeexpectancy.com/peru-alzheimers-dementia

53. Norris ET, Rishishwar L, Chande AT, Conley AB, Ye K, Valderrama-Aguirre A, et al. Admixture-enabled selection for rapid adaptive evolution in the Americas. Genome Biol. 2020;21(1):1–12.

54. Norris ET, Wang L, Conley AB, Rishishwar L, Mariño-Ramírez L, Valderrama-Aguirre A, et al. Genetic ancestry, admixture and health determinants in Latin America. BMC Genomics. 2017;19(861).

55. Casey A. Gifford, Ranade SS, Samarakoon R, Salunga HT, Soysa TY de, Huang Y, et al. Oligogenic inheritance ofa human heart disease involving a genetic modifier. Hum Genet. 2019;364:865–70.

56. Ceder MM, Aggarwal T, Hosseini K, Maturi V, Patil S, Perland E, et al. CG4928 is vital for renal function in fruit flies and membrane potential in cells: A first in-depth characterization of the putative solute carrier UNC93A. Front Cell Dev Biol. 2020;8.

57. Bradfield JP, Qu HQ, Wang K, Zhang H, Sleiman PM, Kim CE, et al. A genome-wide meta-analysis of six type 1 diabetes cohorts identifies multiple associated loci. PLoS Genet. 2011;7(9):e1002293.

58. D’Angelo CS, Varela MC, De Castro CIE, Otto PA, Perez ABA, Lourenço CM, et al. Chromosomal microarray analysis in the genetic evaluation of 279 patients with syndromic obesity. Mol Cytogenet. 2018;11(14).

59. Zenaro E, Piacentino G, Constantin G. The blood-brain barrier in Alzheimer’s disease. Neurobiol Dis [Internet]. 2017;107:41–56.

60. Aguilar-Pineda JA, Vera-Lopez KJ, Shrivastava P, Chávez-Fumagalli MA, Nieto-Montesinos R, Alvarez-Fernandez KL, et al. Vascular smooth muscle cell dysfunction contribute to neuroinflammation and Tau hyperphosphorylation in Alzheimer disease. iScience. 2021;24(9).

61. Zeng K, Yu X, Mahaman YAR, Wang JZ, Liu R, Li Y, et al. Defective mitophagy and the etiopathogenesis of Alzheimer’s disease. Transl Neurodegener. 2022;11(1):1–13.

62. Galati S, Boni C, Gerra MC, Lazzaretti M, Buschini A. Autophagy: a player in response to oxidative stress and DNA damage. Oxid Med Cell Longev. 2019;Article ID.

63. Lall MA, Cox HC, Arcila ML, Cadavid L, Moreno S, Garcia G, et al. Origin of the PSEN1 E280A mutation causing early–onset Alzheimer’s disease. Alzheimers Dement. 2014;10:S277–S283.e10.

64. Tomita T, Maruyama K, Saido TC, Kume H, Shinozaki K, Tokuhiro S, et al. The presenilin 2 mutation (N141I) linked to familial Alzheimer disease (Volga German families) increases the secretion of amyloid β protein ending at the 42nd (or 43rd) residue. Proc Natl Acad Sci U S A. 1997;94(5):2025–30.

